# E-cigarettes With Varenicline Versus Varenicline for Smoking Cessation: A Pragmatic Randomised Controlled Trial

**DOI:** 10.1101/2022.05.16.22275152

**Authors:** Harry Tattan-Birch, Loren Kock, Jamie Brown, Emma Beard, Linda Bauld, Robert West, Lion Shahab

## Abstract

**Background:** We examined whether, in adults receiving behavioural support, offering e-cigarettes together with varenicline helps more people stop smoking cigarettes than varenicline alone.

**Methods:** A two-group, parallel-arm, pragmatic randomised controlled trial was conducted in six English stop smoking services from 2019-2020. Adults enrolled onto a 12-week programme of in-person one-to-one behavioural smoking cessation support (N=92) were randomised to receive either (i) a nicotine e-cigarette starter-kit alongside varenicline or (ii) varenicline alone. The primary outcome was biochemically-verified abstinence from cigarette smoking between weeks nine-to-12 post quit-date, with those lost to follow-up considered not abstinent. The trial was stopped early due to COVID-19 restrictions and a varenicline recall (92/1266 participants recruited).

**Results:** Nine-to-12-week smoking abstinence rates were 47.9% (23/48) in the e-cigarette-varenicline group compared with 31.8% (14/44) in the varenicline-only group, a 51% increase in abstinence among those offered e-cigarettes; however, the confidence interval (CI) was wide, including the possibility of no difference (risk ratio [RR]=1.51, 95%CI=0.91-2.64). The e-cigarette-varenicline group had 43% lower hazards of relapse from continuous abstinence than the varenicline-only group (hazards ratio [HR]=0.57, 95%CI=0.34-0.96). Attendance for 12 weeks was higher in the e-cigarette-varenicline than varenicline-only group (54.2% versus 36.4%; RR=1.49, 95%CI=0.95-2.47), but similar proportions of participants in both groups used varenicline daily for ≥8 weeks after quitting (22.9% versus 22.7%; RR=1.01, 95%CI=0.47-2.20). Estimates were too imprecise to determine how adverse events differed by group.

**Conclusion:** Preliminary evidence suggests offering e-cigarettes alongside varenicline to people receiving behavioural support may be more effective for smoking cessation than varenicline alone.

**Implications:** Offering e-cigarettes to people quitting smoking with varenicline may help them remain abstinent from cigarettes, but the evidence is preliminary because our sample size was smaller than planned — caused by COVID-19 restrictions and a manufacturing recall. This meant our effect estimates were imprecise, and additional evidence is needed to confirm that providing e-cigarettes and varenicline together helps more people remain abstinent than varenicline alone.

## Introduction

Rates of cigarette smoking are declining in many high-income countries,^1^ in part due to the availability of treatments that help people stop smoking.^2^ Varenicline — a partial nicotinic acetylcholine receptor agonist — Is one of the most effective treatments, especially when paired with behavioural support.^3^ However, even with varenicline, fewer than one-in-five people remain abstinent from smoking for a year or more after quitting,^4^ so there remains a need to find more effective options. Electronic cigarettes (e-cigarettes)^5^ have become a popular method of quitting cigarette smoking in England, used in a third of quit attempts.^6^ E-cigarettes can deliver similar amounts of nicotine as cigarettes but, by avoiding tobacco combustion, expose users to much lower levels of toxicants.^7–9^ Offering electronic cigarettes (“e-cigarettes”) alongside varenicline and behavioural support may help people maintain abstinence from smoking conventional cigarettes.

The rationale for providing e-cigarettes alongside varenicline is twofold. First, e-cigarettes mimic the sensory and behavioural aspects of smoking that contribute to dependence,^10^ something which is not provided by varenicline. Second, the pharmacological effects of varenicline may be enhanced by providing additional nicotine. The main target of varenicline, and an important mediator of nicotine dependence,^11^ the α4ß2 nAChR subtype, may not be fully saturated by varenicline, allowing nicotine from other sources to bind to increase receptor activation. Moreover, varenicline does not fully stop the dopaminergic effects of smoking, and additional nicotine may bind to other receptors important to dependence that varenicline does not affect.^12^ It may also be that the pharmacokinetics of varenicline and alternate nicotine-delivery devices complement one another to provide a more favourable agonistic effect on receptors.^13^

Observational data from English stop smoking services show that people who use nicotine e-cigarettes, varenicline, and behavioural support together are more successful in their attempts to quit smoking than those using any other treatment.^14^ Moreover there is trial evidence that combination therapy of nicotine replacement therapy (NRT) and varenicline is safe and well-tolerated and may increase abstinence rates compared with varenicline alone,^12^ particularly for more dependent smokers,^15^ and compared with NRT alone in alcohol-dependent smokers.^16^ However there are no trial data on combination therapy of e-cigarettes with varenicline. E-cigarettes may offer an additional advantage over NRT not only because they more closely mimic cigarettes, but also because they have been found to be more effective nicotine delivery devices, increasing abstinence rates compared with NRT.^17,18^ One ongoing trial in New Zealand is evaluating the effectiveness and safety of combining varenicline with nicotine e-cigarettes for smoking cessation among those with mental health illnesses.^19^ However, as far as we are aware, there are no studies taking place in the general population investigating combination therapy of varenicline with e-cigarettes against varenicline alone in routine stop smoking services. If found to be effective in an RCT, this could become a new gold standard treatment for smoking cessation.

This pragmatic trial aims to answer the following question: in adults receiving one-to-one behavioural support at English stop smoking services, does offering nicotine e-cigarette starter-kits together with varenicline increase cigarette abstinence rates compared with varenicline alone?

We also aim to examine how offering e-cigarettes to clients affects attendance at stop smoking services, adherence to varenicline, and e-cigarette use. Moreover, a qualitative process evaluation aims to explore the acceptability of offering e-cigarettes alongside varenicline at services, as well as barriers and enablers to using them.

## Methods

### Design

This is a two-group, parallel arm, pragmatic randomised controlled trial. It was conducted between April 2019 and March 2020 in stop smoking services in England, which are free to access for smokers trying to quit. Fifteen services were approached to take part in the study, of which eight (53%) agreed to participate and six (40%) started recruitment. Reasons for not participating included lack of staff capacity, incompatible models of service delivery, and concerns about e-cigarettes (Table S1).

Services recruited participants and delivered the intervention during one-to-one in-person counselling sessions with trained stop-smoking advisors. Participants were randomised (1:1 ratio in blocks of 6 or 8 participants, stratified by service) using a computer-generated random sequence with allocation concealed within opaque envelopes. Due to the nature of the intervention, participants and advisors could not be blinded to treatment assignment.

Ethical approval was granted by both University College London (8323/003) and the NHS Health Research Authority (19/LO/0239). The study was overseen by both a trial steering and a data monitoring committee. The trial protocol and analysis plan were registered prior to participant recruitment (ISRCTN16931827) and were peer-reviewed as a registered report at N&TR. Updates were approved by the data monitoring committee prior to unblinding or analysis of data. These updates added secondary analyses of continuous abstinence and respiratory symptoms, as well as sensitivity analyses for the primary outcome (Table S2). The original and updated protocols are available online, alongside a summary of changes (https://osf.io/vm4g3/).

### Procedures

In their first session, smokers were asked to set a target quit date, usually within one to four weeks, and advisors used a checklist to assess eligibility for inclusion in the trial. Cigarette smokers were eligible if they were proficient in English, were not pregnant or breast feeding, opted to use varenicline, were willing to try e-cigarettes, and had not regularly used e-cigarettes in the past six months.

Advisors gave eligible smokers trial information and a consent form. After smokers provided written informed consent, advisors recorded baseline characteristics, took an exhaled carbon monoxide (CO) reading, and opened opaque envelopes to reveal whether smokers were randomised to the e-cigarette-varenicline group or the varenicline-only group.

This study was designed to avoid interfering with standard service protocols. Following existing practice, participants in both randomised groups were prescribed varenicline and given behavioural support during regular in-person sessions with their advisor. They were offered weekly or fortnightly support until 12 weeks after their quit date. Behavioural support aimed to minimise participants’ motivation to smoke, maximise their motivation to remain abstinent, and guide their use of pharmacotherapy — as described in detail elsewhere.^20^ During each session, advisors recorded smoking status, exhaled CO, adherence, adverse events, and respiratory symptoms using existing software (QuitManager or PharmOutcomes).

The COVID-19 pandemic led all in-person sessions to be stopped after March 2020. Advisors remotely followed up with those (n=5) who had yet to complete their final 12-week appointment, using CO-monitors that had been posted to participants to verify abstinence.

#### Varenicline-only group

Participants were prescribed the standard 12-week course of varenicline, starting approximately two weeks prior to their target quit date. They were advised to take one 0.5mg pill daily for the first three days, then two 0.5mg pills daily for days four to seven, and finally two 1mg pills daily for the remaining 11 weeks. As this was a pragmatic trial, participants were not asked to avoid using e-cigarettes.

#### E-cigarette-varenicline group

These participants also received standard 12-week course of varenicline described above. In addition, they were given an e-cigarette starter kit prior to their quit date. The starter kit contained an Aspire PockeX e-cigarette (as used in previous trials),^17,21^ e-liquid to last for approximately four weeks, and an information booklet about e-cigarettes (available here: https://osf.io/59adw/). Participants could choose a total of eight 10ml e-liquid bottles (from Aspire or Totally Wicked) in any combination from a selection of three flavours (fruit, menthol, and tobacco) and three nicotine concentrations (6, 12, and 18mg/ml). Participants were encouraged to buy further bottles from local vape shops. Advisors gave participants brief in-person advice about how to use e-cigarettes and asked them to try the e-cigarette during the session. As this pragmatic trial aimed to test the effect of offering — not using — an e-cigarette, participants were asked but not required to use them.

### Measures

At every session after quitting, participants were asked whether they had smoked cigarettes since their previous session, with exhaled CO-readings of below 10ppm used to verify cigarette abstinence.^22^ They were also asked, since their last session, how frequently they had used varenicline or e-cigarettes and whether they had experienced specific adverse events (sleep disturbance, nausea, throat/mouth irritation) or respiratory symptoms (phlegm, cough, shortness of breath, wheezing). Advisors were required to report serious adverse events to the trial team, but none occurred throughout the trial. Further details about questionnaire items are available in the appendix.

Nine-to-12-week smoking abstinence was the primary outcome, with participants considered abstinent if they (i) reported not smoking cigarettes between weeks nine and 12 after their quit date and (ii) gave a CO-reading below 10ppm at week 12 or later. Participants with missing data for the primary outcome were assumed not to be abstinent.

Secondary abstinence outcomes included two-to-four-week smoking abstinence (defined as above) and length of continuous abstinence. The latter outcome was not included in the original protocol but was added to the updated protocol and registered prior to data analysis (https://osf.io/vm4g3/). It was measured as the number of weeks each participant remained continuously abstinent from smoking before relapsing to smoking.

Attendance was assessed using two outcomes. Firstly, whether or not a participant continued attending sessions until at least 12 weeks after the quit date. Secondly, the number of sessions, of a possible four, a participant attended in their first four weeks after quitting.

Two outcomes assessed adherence to varenicline. Firstly, whether or not participants reported using varenicline daily for at least one week after their quit date and, secondly, whether they used varenicline daily until at least 8-weeks after quitting. The latter allows up to four weeks of varenicline use prior to quitting. E-cigarette outcomes were daily use for at least one week after the quit date and daily use at every session attended after quitting.

Time to first experiencing each adverse event and respiratory symptom were recorded for each participant.

### Analysis

Data analyses were conducted by the trial statistician with blinding to treatment assignments using R version 4.1.3.^23^ Anonymised data and analysis code are openly available (https://osf.io/vdngh/). The primary and other binary outcomes were reported as risk ratios (RR) with 95% confidence intervals (95% CIs). Analyses of binary smoking abstinence outcomes followed the intention-to-treat principle, where all those with missing follow-up data were treated as having relapsed (0% abstinent).

In sensitivity analyses for the primary outcome, RRs were calculated with a range of different assumed abstinence rates in those lost to follow-up (e.g., 10%, 20%, 30% and 40%).

Moreover, for length of continuous abstinence, the hazard ratio (HR) for relapse was estimated using a Cox proportional-hazards model. Participants who were lost to follow-up were assumed to have relapsed in the week after the final stop-smoking session they attended where CO-measurements were taken. Participants who were still abstinent at week 12 were considered censored after this time.

Unplanned sensitivity analyses for the primary outcome adjusted for e-cigarette non-adherence (i.e., people in the e-cigarette-varenicline group who did not try e-cigarettes) and contamination (i.e., people in the varenicline-only group who tried e-cigarettes), using a method described by Cuzick and colleagues.^24^ This provides an estimate of the effect of trying e-cigarettes (daily use for at least a week) among co-operators: individuals who would try e-cigarettes if they were assigned to the e-cigarette-varenicline group, but would not try them if assigned to the varenicline-only group.^25^

Cox models were also used to estimate the HR for time to first experiencing each adverse event and respiratory symptom. These were reported alongside the incidence rate for each randomised group (i.e., number of people who reported an event divided by the person-weeks-at-risk), with the incidence rate ratio (IRR) estimated using a log-rate model. For these analyses, participants were considered censored after the final week they attended a follow-up session (maximum 12 weeks post-quit date).

### Sample size and early stopping

As described in the original study protocol (https://osf.io/vxw8r/), previous literature suggested an expected risk ratio of 1.26 for our primary outcome.^12,14^ It was determined that a sample of 633 participants per group would provide at least 90% power to detect this effect size in a two-tailed analysis.

Restrictions introduced in response to the COVID-19 pandemic caused services to move sessions online, which meant advisors could not provide e-cigarettes to participants or take in-person CO-readings. This led the trial to be paused in March 2020, before the target number of participants had been recruited (92/1266). Plans to run the trial remotely were halted when, in July 2021, Pfizer recalled Champix (the only form of varenicline available in England) due to levels of N-nitroso-varenicline that were higher than considered acceptable by the European Medicines Agency.^26^ In agreement with the funder, Pfizer, the trial was stopped in November 2021.

### Process evaluation

Quantitative process evaluation included summaries of attendance at stop smoking services, varenicline adherence, and e-cigarettes adherence/contamination.

Qualitative process evaluation involved semi-structured interviews using a flexible topic guide (https://osf.io/2pgz4/), carried out with ten participants from the e-cigarette-varenicline group who had been followed up until at least four weeks after their quit date. Interview transcripts were analysed in two stages, using a combination of deductive and inductive thematic framework analysis. Firstly, themes surrounding the acceptability of services providing e-cigarettes alongside varenicline were classified under the theoretical framework of acceptability (TFA).^27^ Then, barriers and enablers to using e-cigarettes for smoking cessation under the COM-B model were identified.^28^ More details about the process evaluation and are provided elsewhere.^29^

## Results

### Participants

Of the 92 cigarette smokers randomised at stop-smoking services between April 2019 and March 2020, 48 were assigned to the e-cigarette-varenicline group and 44 to the varenicline-only group. Participants had a mean age of 43.9 (SD=13.1), 51% (n=47) were women, 79% (n=73) were white, and 29% (n=27) had routine or manual occupations (Table 1). Table 1 shows that participants in both randomised groups had similar baseline characteristics. Of those randomised, 46% (n=42) attended follow-up sessions for at least 12 weeks after their quit date (Figure 1).

**Table 1:** Baseline characteristics*.

|  | **E-cigarette** | **Control** | **Combined** |
| --- | --- | --- | --- |
| N | 48 | 44 | 92 |
| Age | 43.8 ± 12.1 | 44.0 ± 14.2 | 43.9 ± 13.1 |
| Gender |  |  |  |
| Woman | 52% (25) | 50% (22) | 51% (47) |
| Man | 48% (23) | 50% (22) | 49% (45) |
| Ethnicity |  |  |  |
| White | 79% (38) | 80% (35) | 79% (73) |
| Black or Asian | 17% (8) | 11% (5) | 14% (13) |
| Other or mixed | 4% (2) | 9% (4) | 7% (6) |
| Occupation |  |  |  |
| Managerial or professional | 40% (19) | 39% (17) | 39% (36) |
| Routine or manual | 27% (13) | 32% (14) | 29% (27) |
| Other† | 33% (16) | 30% (13) | 32% (29) |
| Free prescription |  |  |  |
| Not reported | 71% (34) | 66% (29) | 68% (63) |
| Yes | 29% (14) | 34% (15) | 32% (29) |
| Anxious or depressed |  |  |  |
| No | 77% (37) | 68% (30) | 73% (67) |
| Yes | 24% (11) | 32% (14) | 27% (25) |
| Cigarettes per day‡ |  |  |  |
| ≤10 | 15% (3) | 30% (7) | 23% (10) |
| 11-20 | 45% (9) | 48% (11) | 47% (20) |
| 21-30 | 30% (6) | 22% (5) | 26% (11) |
| ≥31 | 10% (2) | 0% (0) | 5% (2) |

\* Age presented as mean ± standard deviation. All other characteristics summarised as % (n).
† Includes people who are retired, unemployed or home carers.
‡ Only recorded for 43 participants: 20 in the e-cigarette-varenicline (e-cigarette) group and 23 in the varenicline-only (control) group.

**Figure 1.**
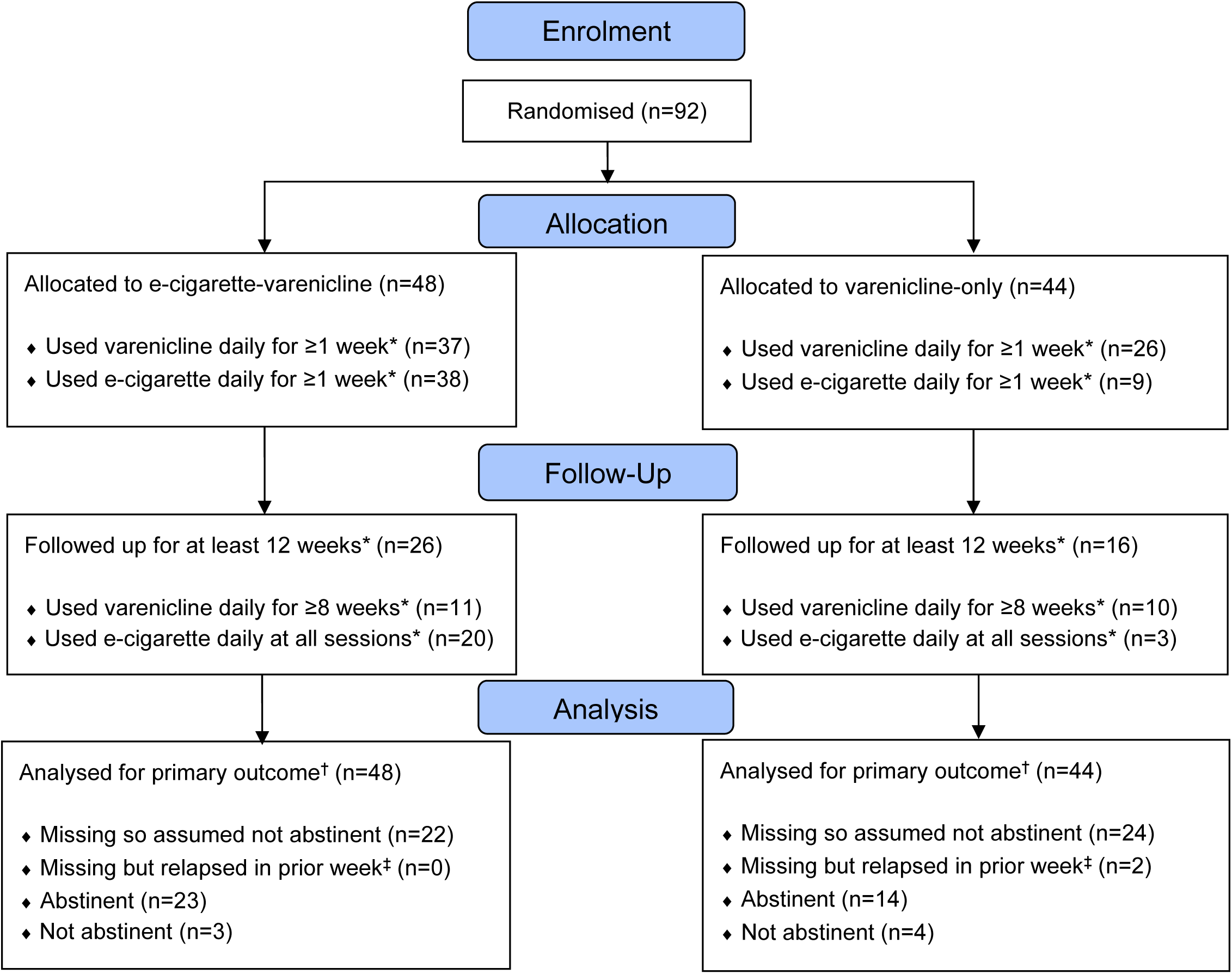
CONSORT flow diagram. A software issue meant it was only possible to determine the number of participants who were both eligible for and willing to take part in the trial, not the total number who were approached. Reasons for loss to follow-up were not recorded due to the pragmatic nature of the trial. * After their quit date. † Nine-to-12-weeks abstinence from cigarette smoking, biochemically verified with exhaled CO under 10ppm. ‡ Missing between weeks nine and twelve but reported relapse prior to week nine.

### Smoking abstinence

#### Primary — Nine-to-12-week abstinence

Nine-to-12-week abstinence rates were 47.9% (n=23) in the e-cigarette-varenicline group compared with 31.8% (n=14) in the varenicline-only group. This equates to a 1.51-fold increase in abstinence rates in those offered e-cigarettes; however, the confidence interval was wide and included the possibility of no difference (RR 1.51, 95%CI 0.91-2.64). Bayes factors are shown in Table S3 Results were similar when including quits that were self-reported but not biochemically verified (52.1% versus 34.1%; RR 1.53, 95%CI 0.95-2.60).

Table S4 shows sensitivity analyses which relax the assumption that all participants missing for the follow-up had relapsed. These show that the higher the percentage of missing participants who were abstinent, the smaller the estimated effect size (e.g., RR 1.38 if 20% of missing participants were abstinent).

#### Secondary — Two-to-four-week abstinence

Two-to-four-week abstinence rates were 1.37 times higher in the e-cigarette-varenicline than varenicline-only group, but the confidence interval was compatible with effects ranging from just under no difference to 2.01 times higher rates in those offered e-cigarettes (68.8% versus 50.0%; RR 1.37, 95%CI 0.98-2.01).

#### Secondary — Continuous abstinence

The e-cigarette-varenicline group had a 43% lower (instantaneous) rate of relapse to cigarette smoking than those in the varenicline-only group (Cox model; HR 0.57, 95%CI 0.34-0.96). Figure 2 shows a Kaplan-Meier plot for the length of time each participant remained continuously abstinent from cigarettes before relapsing. Note that these analyses were not included in the original protocol but were added to the updated protocol which was registered prior to data analysis.

**Figure 2.**
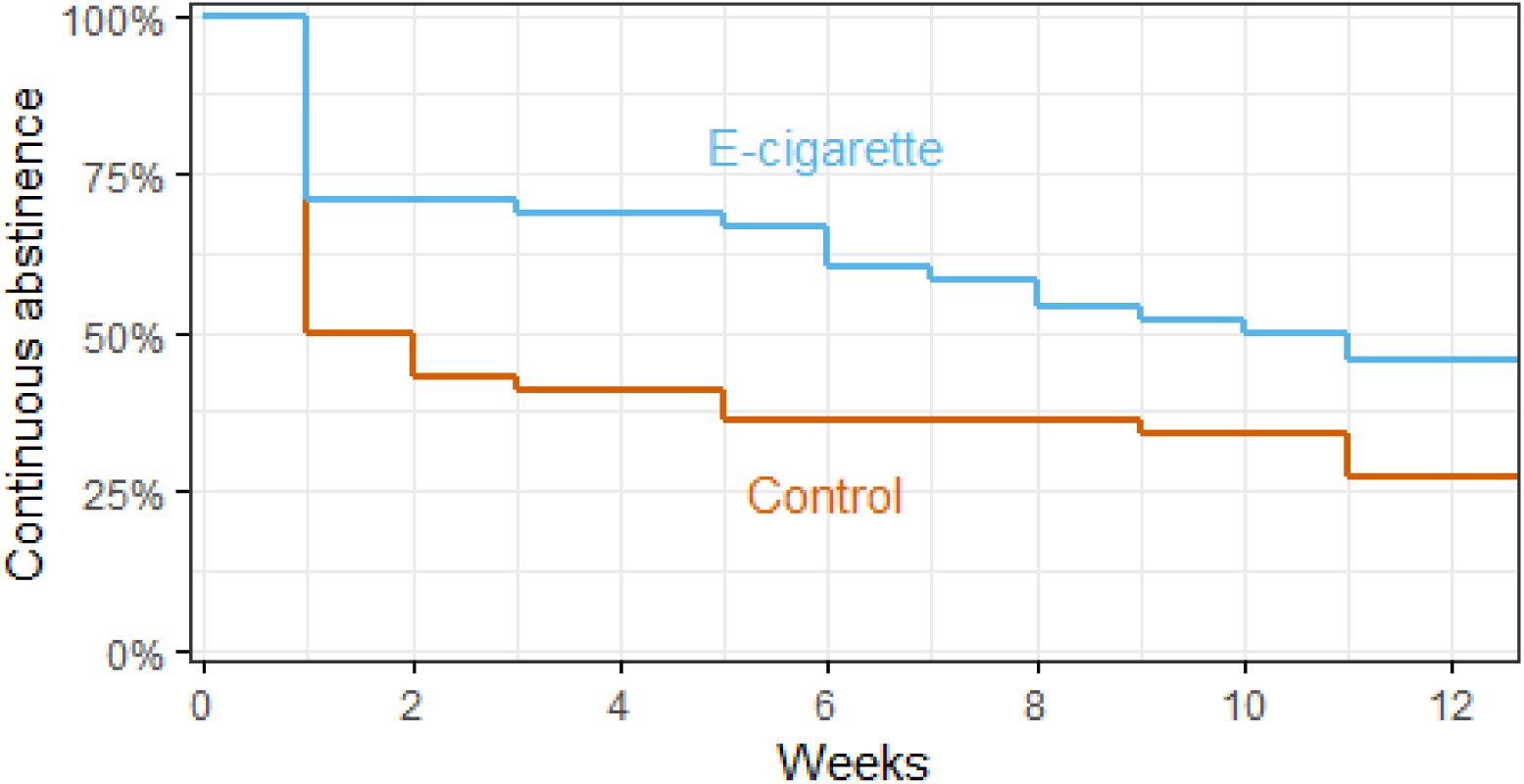
Kaplan-Meier plot showing the percentage of participants continuously abstinent (CO<10ppm) from cigarette smoking at each week after their quit date. Participants who were lost to follow-up were assumed to have relapsed in the week after the final session they attended.

### Safety

#### Adverse events

Overall, 59.8% (n=55) of participants experienced at least one adverse event between their quit date and final session. Sleep disturbance was reported by 44.6% (n=41) of participants, nausea by 34.8% (n=32), and throat or mouth irritation by 27.2% (n=25). Comparisons of event incidence rates and hazard ratios between the e-cigarette-varenicline and varenicline-only group are shown in Table 2. These estimates were too imprecise to determine the size or direction of differences between groups (e.g., any adverse event; HR 0.69, 95%CI 0.40-1.20). Risks of adverse events among those followed-up for at least 12 weeks are shown in Table S5. No serious adverse events were reported in either group.

**Table 2:** Incidence of adverse event and respiratory symptoms.

|  | Group* | Events | Weeks† | Rate‡ | IRR (95%CI)‡ | HR (95%CI)‡ |
| --- | --- | --- | --- | --- | --- | --- |
| **Adverse events** |  |  |  |  |  |  |
| Any | Control | 24 | 144 | 1.67 | Ref | Ref |
|  | E-cigarette | 31 | 193 | 1.61 | 0.96 (0.57-1.66) | 0.69 (0.40-1.20) |
| Sleep disturbance | Control | 21 | 163 | 1.29 | Ref | Ref |
|  | E-cigarette | 20 | 283 | 0.71 | 0.55 (0.30-1.02) | 0.64 (0.34-1.20) |
| Nausea | Control | 14 | 209 | 0.67 | Ref | Ref |
|  | E-cigarette | 18 | 261 | 0.69 | 1.03 (0.51-2.11) | 0.84 (0.41-1.72) |
| Throat/mouth irritation | Control | 7 | 244 | 0.29 | Ref | Ref |
|  | E-cigarette | 18 | 310 | 0.58 | 2.02 (0.88-5.21) | 1.11 (0.45-2.74) |
| **Respiratory symptoms** |  |  |  |  |  |  |
| Any | Control | 19 | 164 | 1.16 | Ref | Ref |
|  | E-cigarette | 25 | 258 | 0.97 | 0.84 (0.46-1.54) | 1.05 (0.57-1.92) |
| Phlegm | Control | 13 | 210 | 0.62 | Ref | Ref |
|  | E-cigarette | 20 | 293 | 0.68 | 1.10 (0.55-2.27) | 0.75 (0.37-1.53) |
| Cough | Control | 14 | 200 | 0.70 | Ref | Ref |
|  | E-cigarette | 17 | 301 | 0.56 | 0.81 (0.40-1.66) | 1.49 (0.72-3.08) |
| Shortness of breath | Control | 8 | 225 | 0.36 | Ref | Ref |
|  | E-cigarette | 12 | 332 | 0.36 | 1.02 (0.42-2.59) | 1.22 (0.48-3.10) |
| Wheezing | Control | 6 | 249 | 0.24 | Ref | Ref |
|  | E-cigarette | 7 | 381 | 0.18 | 0.76 (0.25-2.37) | 0.85 (0.26-2.82) |

\* There were 44 participants in the varenicline-only (control) group and 48 in the e-cigarette-varenicline (e-cigarette) group.
† Total person-weeks at risk of first event. For each person, this is the number of weeks from the quit date until they either experienced the event/symptom, were lost to follow-up, or completed the study (12 weeks post-quit).
‡ Incidence rate calculated per 10 person-weeks. Incidence rate ratios (IRR) and corresponding 95% confidence intervals (95%CI) estimated using log-linear rate models. Hazards ratios (HR) and corresponding 95% CIs estimated using Cox proportional-hazards models. Schoenfeld tests found some evidence for non-proportional hazards for throat/mouth irritation ( $p=.046$ ) and cough ( $p=.032$ ), but all other outcomes were compatible with proportionality ( $p>.31$ ).

#### Respiratory symptoms

Respiratory symptoms were reported by 47.8% (n=44) of participants at least once between their quit date and the final session they attended. Phlegm was reported by 35.9% (n=33) of participants, cough by 33.7% (n=31), shortness of breath by 21.7% (n=20), and wheezing by 14.1% (n=13). Table 2 shows that rates of respiratory symptoms were similar in the e-cigarette-varenicline and varenicline-only group (e.g., any symptom; HR 1.05, 95%CI 0.57-1.92), but confidence intervals included the possibility of meaningful differences between groups.

### Process evaluation: Quantitative data

#### Attendance

Of the 92 participants randomised, 45.7% (n=42) continued attending stop-smoking service sessions for at least 12 weeks after their quit date. Attendance for 12 weeks was 54.2% (n=26) in the e-cigarette-varenicline group compared with 36.4% (n=16) in the varenicline-only group (RR 1.49, 95%CI 0.95-2.47). On average, participants in the e-cigarette-varenicline group attended 3.1 out of a possible four sessions in the first four weeks after quitting, while those in the varenicline-only group attended 2.8 sessions (proportional-odds model; OR 1.69, 95%CI 0.93-2.45).

#### Varenicline adherence

In the e-cigarette-varenicline group, 77.1% (n=37) of participants used varenicline daily for at least one week after their quit date, compared with 59.1% (n=26) in the varenicline-only group (RR 1.30, 95%CI 0.99-1.79). Daily varenicline use for at least eight weeks after quitting was reported by 22.9% (n=11) of participants in the e-cigarette-varenicline group and 22.7% (n=10) in the varenicline-only group (RR 1.01, 95%CI 0.47-2.20).

#### E-cigarette adherence and contamination

In the e-cigarette-varenicline group, 79.2% (n=38) used e-cigarettes daily for at least one week after their quit date, and 41.7% (n=20) reported daily use at every session they attended after quitting. There was some contamination: 20.5% (n=9) of participants in the varenicline-only group used e-cigarettes daily for at least one week after their quit date, and 6.8% (n=3) reported daily use at every session they attended after quitting.

In an unplanned analysis of the primary outcome that adjusted for non-adherence (i.e., being assigned to try e-cigarettes but not doing so) and contamination (i.e., being assigned to the control group but trying e-cigarettes), trying e-cigarettes was estimated to increase nine-to-12-week abstinence by 2.66 times (RR 2.66, 96%CI 1.17-6.05).^24^

### Process evaluation: Qualitative data

#### Acceptability

Themes surrounding the acceptability of providing e-cigarettes alongside varenicline at stop smoking services were identified from semi-structured interviews with ten participants in the e-cigarette-varenicline group (Table S6 and https://osf.io/2pgz4/). Participants perceived the intervention package as complementary, with varenicline reducing urges to smoke and the e-cigarette replacing the habit of smoking. However, some were concerned that e-cigarettes may ‘replace one addiction with another’ and, thus, there were mixed opinions about whether services should provide e-cigarette.

#### Barriers and enablers

Barriers and enablers to using e-cigarettes for smoking cessation are shown in Table S7. Enablers included the perception that e-cigarettes replace the habit of smoking and offer a ‘back-up’ to varenicline and behavioural support when participants are most at risk of relapse. E-cigarettes were also described as cheaper than cigarettes and could be used in more situations than smoking. Some participants reported the harshness of puffing of e-cigarettes as a barrier to using them, especially early in their quit attempt.

## Discussion

### Summary

Our study provides preliminary evidence that, among people receiving one-to-one behavioural support, offering e-cigarettes alongside varenicline may be more effective for cigarette smoking cessation than varenicline alone. The evidence is preliminary because our sample size was smaller than planned — caused by COVID-19 and a manufacturing recall — which meant our effect estimates were imprecise (highly compatible with 9% lower to 164% greater nine-to-12-week abstinence rates in those given e-cigarettes). More data are needed to confirm whether providing e-cigarettes and varenicline together help more people remain abstinent than varenicline alone.

### Comparison with prior literature

Nonetheless, our study adds to a wider literature on the effects of offering alternative nicotine products alongside varenicline. Our results closely align with a previous meta-analysis finding the 50% higher odds of cigarette abstinence in those given NRT alongside varenicline than varenicline alone (OR 1.50, 95%CI 1.14-1.97).^12^ However, another recent study showed that adding nicotine patches to varenicline had little effect on abstinence rates (OR 0.99, 95% CI 0.87-1.12).^30^ It is possible that fast acting nicotine products — including gums, sprays, e-cigarettes — are better at helping varenicline users remain abstinent, as they can satisfy momentary urges for nicotine.^31^

### Interpretation

We found that most of the difference in relapse between groups occurred within the first week after quitting. Three quarters of participants in the e-cigarette group remained abstinent for at least one week compared with just half of those in the varenicline-only group (Figure 2). This could be explained by e-cigarettes helping people overcome the intense urges to smoke most people experience in the first few days after quitting.^32–34^ However, it is possible that some people entered the study because they wanted a free e-cigarette. In learning they had been randomised to the control group, they may have bought an e-cigarette elsewhere and stopped attending sessions. Because the primary analysis assumed that people with missing follow-up data had relapsed to smoking, this could lead us to overestimate differences in abstinence rates between groups (as shown in sensitivity analyses in Table S1).

Conversely, other factors could have led us to underestimate any effect using e-cigarettes had on abstinence. For instance, there was some non-adherence with only 79% of participants in the e-cigarette-varenicline group trying their e-cigarette for at least a week after quitting. There was also some contamination, with 20% of those in the varenicline-only group trying e-cigarettes. In unplanned sensitivity analyses adjusting for this non-adherence and contamination, our estimate for the effect of e-cigarettes on increasing nine-to-12-week abstinence tripled from 51% to 166%.

Secondary analyses indicated that e-cigarettes help people remain continuously abstinent from cigarettes for longer, with data most compatible with 43% lower instantaneous rate of relapse in the e-cigarette-varenicline than varenicline-only group. This analysis rested on the assumption that people who were lost to follow-up had relapsed in the week after the final session they attended. However, because lost to follow-up was greater in the varenicline-only group, this could have biased results in favour of e-cigarettes.

In interviews, participants reported that they viewed the e-cigarettes, varenicline and behavioural support to be acceptable and complementary, but some were concerned about continued nicotine use and the harshness of vaping. These concerns may be alleviated by providing information around the relative harms of smoking versus vaping,^35^ giving advice about titrating inhalation to avoid harshness, or providing products that are less harsh to inhale — such as those using lower pH nicotine salts e-liquid.^36^

### Strengths and limitations

The study benefits from using randomised assignment, which provides internal validity (exchangeability), and a pragmatic design within stop smoking services that guarantees ecological validity (given that this is the setting where such an intervention would likely be implemented). However, there were several limitations. First, clients could not be blinded to their assigned group. This is an inherent limitation of many smoking cessation trials. We partially militated against it by using objective biochemical measures (CO-readings) to verify abstinence from cigarette smoking, which reduces the risk of outcome assessment being biased by assessors knowing which group participants were assigned to. Second, services only followed up clients for 12 weeks after quitting and, because this is a pragmatic trial, we did not ask them to extend this period. This meant abstinence was measured for less than the six months recommended by Russell Standard guidelines.^37^ Third, just under half of participants continued attending services until their final 12 week follow-up session, with 50% greater lost to follow-up in the e-cigarette-varenicline than varenicline-only group. Our primary analysis assumed those with missing follow-up data had relapsed, which is likely a reasonable assumption as people tend to only continue attending services if they remain abstinent. Nonetheless, in sensitivity analyses (Table S4) we quantitatively assessed how certain violations of this assumption would affect results.^38^ Forth, a fifth of those in the varenicline-only group used e-cigarettes while a fifth of those in the e-cigarette-varenicline group did not. This contamination and non-adherence would dilute any effect of using e-cigarettes on abstinence, but we accounted for this in a sensitivity analysis. Finally, trial recruitment was stopped early due to the COVID-19 pandemic and recall of varenicline by Pfizer, which meant we lacked a sufficiently large sample to precisely estimate effects of treatment.

### Conclusion

In conclusion, we found preliminary evidence that, among people receiving one-to-one behavioural support, providing e-cigarettes alongside varenicline may be more effective than offering varenicline alone. However, estimates were imprecise due to the lower than planned sample size; for the primary outcome, anything from 9% lower to 164% higher abstinence rates remained highly compatible with the data (at the 95% level). More data are needed to clarify the effect of adding e-cigarettes to smoking cessation treatment with varenicline.

## Data Availability

Anonymised data and analysis code are openly available (https://osf.io/vdngh/).

https://osf.io/vm4g3/

## Additional information

### Author contributions statement

LS conceived the original idea for this study. LS, LB, JB and RW obtained funding. HTB and LK wrote the initial draft with further input from LS, LB, JB and RW. LS is guarantor for this article. All authors read, reviewed and approved the final version. All researchers listed as authors are independent from the funders and all final decisions about the research were taken without constraint by the investigators. LS had final responsibility for the decision to submit for publication.

## Acknowledgements

We thank advisors at NHS stop smoking services in Bexley, Lewisham, Hackney, Tower Hamlets, Cambridge, and Waltham Forest, who recruited, advised and followed up with participants in the trial. We also thank members of the trial steering committee (Prof Michael Ussher, Dr Dunja Przulj, Prof Peter Hajek, and Dr Felix Naughton) for overseeing and advising on the conduct of the trial. Thanks also go to the members of the data monitoring committee (Dr Natalie Walker, Prof Hayden McRobbie, Dr Leonie Brose, Prof Marcus Munafo) for reviewing the collection, management and analysis of data for the trial. We would also like to acknowledge Dr Fabiana Lorencatto for valuable support with the development and analysis of components in the process evaluation. Finally, thank you to the team at Oxford University who provided us with a template for an information booklet about e-cigarettes, which we adapted and gave to participants.

## Supplementary Tables and Figures

**Table S1:** Reasons for stop smoking services not participating in the trial.

| Service* | Reason for not participating |
| --- | --- |
| 1 | No response after initial contact |
| 2 | Delivery of services through pharmacies incompatible with trial procedures |
| 3 | Service closed down before the trial started |
| 4 | Lack of staff capacity |
| 5 | Delivery of services through general practices incompatible with trial procedures |
| 6 | No response after initial contact |
| 7 | Perceived lack of evidence on e-cigarette harms and use for smoking cessation |

\* Names of services are excluded for data protection purposes

**Table S2:** Summary of planned and unplanned analyses.*

| **Planned and registered before data collection** | **Planned/updated and registered before data analysis** | **Unplanned** |
| --- | --- | --- |
| Analyses of smoking-related outcomes following intention-to-treat principle where those lost to follow-up are treated as smokers | Sensitivity analyses for the primary outcome where risk ratios were calculated with a range of different assumed abstinence rates in those lost to follow-up | Sensitivity analysis for the primary outcome adjusting for e-cigarette non-adherence and contamination |
| Bayes factors for the primary outcome | Hazard ratio (HR) for relapse from continuous abstinence estimated using a Cox model |  |
| Treatment adherence (varenicline adherence and e-cigarette use) across groups | HR and incidence rate ratio for adverse events and respiratory symptoms in the e-cigarette versus control group |  |
| Interviews with ten participants in e-cigarette arm on acceptability and barriers and enablers to participation | Attendance at stop smoking services across groups |  |

\* Reasons for updates to the protocol are discussed in detail online (https://osf.io/vm4g3/).

**Table S7:**
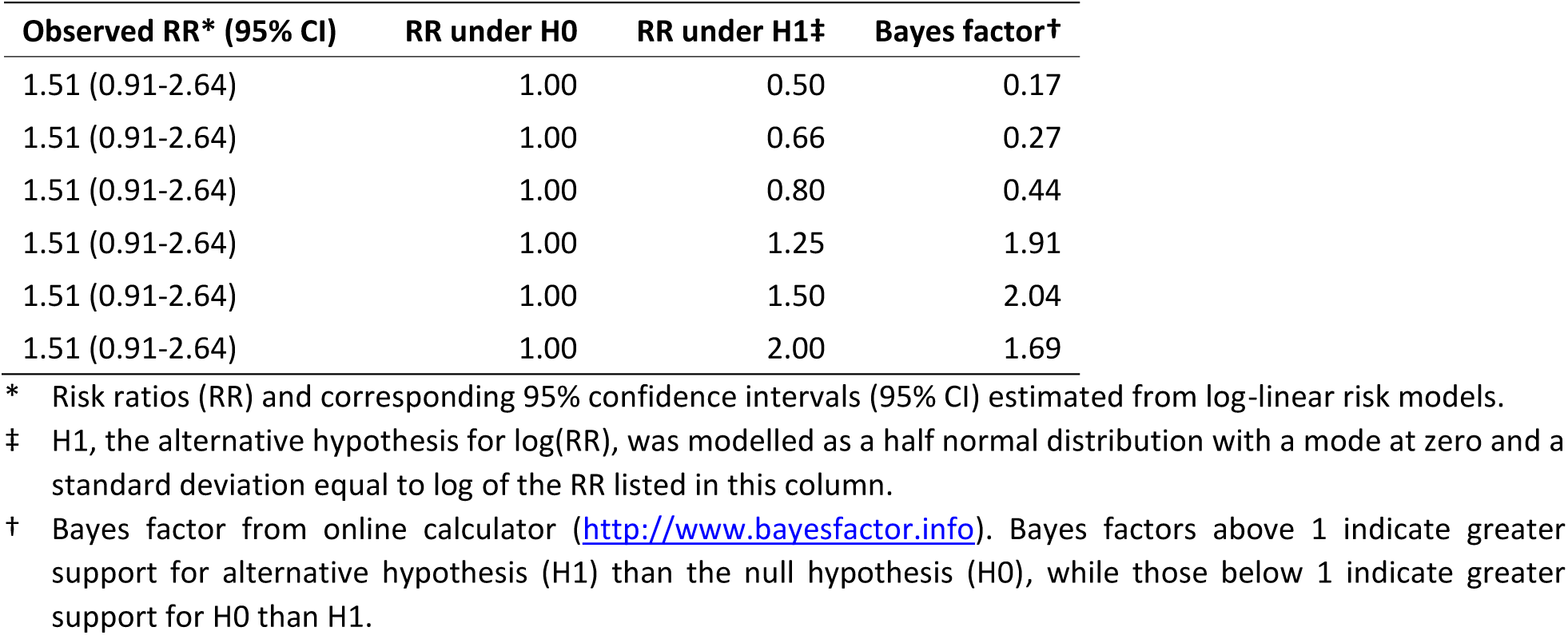
Bayes factors calculated for the primary outcome, nine-to-12 weeks cigarette abstinence.

**Table S4:** Nine-to-12-week cigarette CO-verified abstinence rates when relaxing the assumption that participants with missing follow-up data at week 12 had relapsed (i.e., 0% abstinence rate).

| **Imputed abstinence rate in missing\*** | **Group** | **Missing / N** | **Abstinence rate (n)†** | **RR†** |
| --- | --- | --- | --- | --- |
| 0% | Control | 28 / 44 | 31.8% (14.0) | Ref |
|  | E-cigarette | 22 / 48 | 47.9% (23.0) | 1.51 |
| 10% | Control | 28 / 44 | 36.8% (16.2) | Ref |
|  | E-cigarette | 22 / 48 | 50.8% (24.4) | 1.38 |
| 20% | Control | 28 / 44 | 41.8% (18.4) | Ref |
|  | E-cigarette | 22 / 48 | 53.8% (25.8) | 1.29 |
| 30% | Control | 28 / 44 | 46.8% (20.6) | Ref |
|  | E-cigarette | 22 / 48 | 56.7% (27.2) | 1.21 |
| 40% | Control | 28 / 44 | 51.8% (22.8) | Ref |
|  | E-cigarette | 22 / 48 | 59.6% (28.6) | 1.15 |

\* Imputed abstinence rate among participants who were missing at the 12 weeks post-quit follow-up appointment.
† Estimated percentage and number (n) of people abstinent from cigarette smoking between weeks nine and 12 post-quit, after imputing the abstinence rate in those missing at follow-up. Risk ratio (RR) calculated from these estimates.

**Table S5:** Adverse event risk among those attending the week 12 follow-up session.

| Adverse event | Group | Events* | N | Risk | RR (95%CI)† |
| --- | --- | --- | --- | --- | --- |
| Any | Control | 12 | 16 | 75.0% | Ref |
|  | E-cigarette | 21 | 26 | 80.8% | 1.08 (0.77-1.51) |
| Sleep disturbance | Control | 11 | 16 | 68.8% | Ref |
|  | E-cigarette | 14 | 26 | 53.8% | 0.78 (0.48-1.27) |
| Nausea | Control | 6 | 16 | 37.5% | Ref |
|  | E-cigarette | 14 | 26 | 53.8% | 1.44 (0.69-2.97) |
| Throat/mouth irritation | Control | 6 | 16 | 37.5% | Ref |
|  | E-cigarette | 13 | 26 | 50.0% | 1.33 (0.64-2.80) |

\* Number of participants experiencing at least one event between their quit date and their final follow-up session.
† Risk ratios (RR) and corresponding 95% confidence intervals (95% CI) estimated from log-linear risk models.

**Table S6:** Summary of findings on acceptability of the intervention.

| **TFA domain** | **Theme** | **Effect on acceptability\*** |
| --- | --- | --- |
| Affective attitude | Positive affect for advisor | + |
| Burden | Difficulties with service care pathway | - |
|  | Side-effects from varenicline | - |
| Ethicality | E-cigarette replaces one addiction with another | - |
|  | Opinions about services providing e-cigarettes | +/- |
| Intervention coherence | Complementary nature of intervention package | + |
| Perceived effectiveness | Varenicline reduces urges to smoke | + |

\* + = enhances acceptability; - = reduces acceptability; +/- = differing effects on acceptability. See supporting data at https://osf.io/2pgz4/.

**Table S7:** Summary of findings on barriers and enablers to using e-cigarettes for smoking cessation

| COM-B domain | Theme | Barrier or Enabler * |
| --- | --- | --- |
| Automatic motivation | Replacing the habit of smoking | E |
| Reflective motivation | E-cigarette as a back-up in the quit attempt | E |
|  | E-cigarette is a short-term tool to quit smoking | E/M |
| Physical capability | Harshness of puffing | B |
| Physical opportunity | Cost saving | E |
|  | Opportunities to vape | E |
| Social opportunity | Family support to quit smoking | E |

\* B = barrier; E = enabler; M = mixed. See supporting data at https://osf.io/2pgz4/.

## Appendix

### Questions added to data collection system at services

#### Trial eligibility

Eligible participant agreed to participate in UCL trial (Y/N)

#### Trial arm allocation

If Y selected above, enter treatment allocation (E-cigarette/Control)

#### Varenicline adherence

How often have used Varenicline since last session?

- N/A
- Daily
- Weekly
- Less Than Weekly
- Did not use

#### E-cigarette usage

If the e-cigarette checkbox is checked a further two fields will appear:

- Date device given
- Date field with calendar helper (will retain date from previous session if already populated)

How often have used e-cigarette since last session?

- Not Applicable
- Daily
- Weekly
- Less Than Weekly
- Did not use

#### Adverse reactions

Since the last visit/contact, has the participant experienced any of the following adverse reactions:

- Nausea (Y/N)
- Sleep disturbance (Y/N)
- Throat or mouth irritation (Y/N)

#### Respiratory symptoms

Since the last visit/contact, has the participant experienced any of the following respiratory symptoms:

- Shortness of breath (Y/N)
- Wheezing (Y/N)
- Cough (Y/N)
- Phlegm (Y/N)

#### Mental health

Please select one of the below that describes the participant’s health TODAY:

- not anxious or depressed
- slightly anxious or depressed
- moderately anxious or depressed
- severely anxious or depressed
- extremely anxious or depressed

**CONSORT 2010 checklist of information to include when reporting a randomised trial\***

| Section/Topic | Item No | Checklist item | Reported on page No |
| --- | --- | --- | --- |
| **Title and abstract** |  |  |  |
|  | 1a | Identification as a randomised trial in the title | 1 |
|  | 1b | Structured summary of trial design, methods, results, and conclusions (for specific guidance see CONSORT for abstracts) | 1 |
| **Introduction** |  |  |  |
| Background and objectives | 2a | Scientific background and explanation of rationale | 3,4 |
|  | 2b | Specific objectives or hypotheses | 3,4 |
| **Methods** |  |  |  |
| Trial design | 3a | Description of trial design (such as parallel, factorial) including allocation ratio | 5 |
|  | 3b | Important changes to methods after trial commencement (such as eligibility criteria), with reasons | 6,7,8 |
| Participants | 4a | Eligibility criteria for participants | 5 |
|  | 4b | Settings and locations where the data were collected | 5 |
| Interventions | 5 | The interventions for each group with sufficient details to allow replication, including how and when they were actually administered | 5,6 |
| Outcomes | 6a | Completely defined pre-specified primary and secondary outcome measures, including how and when they were assessed | 6,7 |
|  | 6b | Any changes to trial outcomes after the trial commenced, with reasons | 7 |
| Sample size | 7a | How sample size was determined | 8 |
|  | 7b | When applicable, explanation of any interim analyses and stopping guidelines | N/A |
| Randomisation:<br>Sequence generation | 8a | Method used to generate the random allocation sequence | 5 |
|  | 8b | Type of randomisation; details of any restriction (such as blocking and block size) | 5 |
| Allocation concealment mechanism | 9 | Mechanism used to implement the random allocation sequence (such as sequentially numbered containers), describing any steps taken to conceal the sequence until interventions were assigned | 5 |
| Implementation | 10 | Who generated the random allocation sequence, who enrolled participants, and who assigned participants to interventions | 5 |
| Blinding | 11a | If done, who was blinded after assignment to interventions (for example, participants, care providers, those assessing outcomes) and how | N/A |
|  | 11b | If relevant, description of the similarity of interventions | 6 |
| Statistical methods | 12a | Statistical methods used to compare groups for primary and secondary outcomes | 7 |
|  | 12b | Methods for additional analyses, such as subgroup analyses and adjusted analyses | 7 |
| **Results** |  |  |  |
| Participant flow (a diagram is strongly recommended) | 13a | For each group, the numbers of participants who were randomly assigned, received intended treatment, and were analysed for the primary outcome | 9 |
|  | 13b | For each group, losses and exclusions after randomisation, together with reasons | 9 |
| Recruitment | 14a | Dates defining the periods of recruitment and follow-up | 9 |
|  | 14b | Why the trial ended or was stopped | 6 |
| Baseline data | 15 | A table showing baseline demographic and clinical characteristics for each group | 14 |
| Numbers analysed | 16 | For each group, number of participants (denominator) included in each analysis and whether the analysis was by original assigned groups | 7 |
| Outcomes and estimation | 17a | For each primary and secondary outcome, results for each group, and the estimated effect size and its precision (such as 95% confidence interval) | 9,10,11 |
|  | 17b | For binary outcomes, presentation of both absolute and relative effect sizes is recommended | 9 |
| Ancillary analyses | 18 | Results of any other analyses performed, including subgroup analyses and adjusted analyses, distinguishing pre-specified from exploratory | 9,10,11 |
| Harms | 19 | All important harms or unintended effects in each group (for specific guidance see CONSORT for harms) | 10, 11 |
| **Discussion** |  |  |  |
| Limitations | 20 | Trial limitations, addressing sources of potential bias, imprecision, and, if relevant, multiplicity of analyses | 13 |
| Generalisability | 21 | Generalisability (external validity, applicability) of the trial findings | 12, 13 |
| Interpretation | 22 | Interpretation consistent with results, balancing benefits and harms, and considering other relevant evidence | 12, 13 |
| **Other information** |  |  |  |
| Registration | 23 | Registration number and name of trial registry | 5 |
| Protocol | 24 | Where the full trial protocol can be accessed, if available | 5 |
| Funding | 25 | Sources of funding and other support (such as supply of drugs), role of funders | 13 |

\*We strongly recommend reading this statement in conjunction with the CONSORT 2010 Explanation and Elaboration for important clarifications on all the items. If relevant, we also recommend reading CONSORT extensions for cluster randomised trials, non-inferiority and equivalence trials, non-pharmacological treatments, herbal interventions, and pragmatic trials. Additional extensions are forthcoming: for those and for up to date references relevant to this checklist, see www.consort-statement.org.

